# Genome-wide association study reveals genetic risk factors for trigeminal neuralgia

**DOI:** 10.1101/2021.02.08.21251349

**Authors:** Andrew T. Hale, Jing He, Oluwatoyin Akinnusotu, Rebecca L. Sale, Janey Wang, Lisa Bastarache, Eric R. Gamazon

## Abstract

**Background:** While many clinical risk factors of trigeminal neuralgia (TN) have been identified, the genetic basis of TN is largely unknown. Here, we perform the first genome-wide association study (GWAS) for TN using three independent DNA biobanks – BioVU, the UK Biobank, and Finngen.

**Objective:** To elucidate the genetic basis of TN.

**Methods:** Using GWAS summary statistics generated from BioVU, the UK Biobank, and Finngen, we performed fixed-effect meta-analysis across 490,912 individuals (1,188 TN cases and 489,724 controls) to identify genetic risk factors for TN. Genome-wide significance was defined as p < 5.0×10^−8^.

**Results:** We identify an intergenic locus on chromosome 1p22.2 flanked by *ZNF326* and *SNORD3G* containing 5 SNPs (rs77449572, rs543311093, rs35117749, rs71666259, and rs116010656) reaching genome-wide significance (p < 5.0 x 10^−8^), where rs77449572 is the sentinel variant (p = 1.72 x 10^−9^). The SNP rs77449572 overlaps an enhancer element in cortex-derived neurospheres. In addition, rs71666259 and rs116010656 are located in enhancer elements in embryonic stem cells (HUES48), suggesting potential functional consequences of this locus. We also identify a second locus on chromosome 5q35.1 containing sentinel variant rs62376947 reaching genome-wide significance (p = 2.49 x 10^−8^).

**Conclusions:** To our knowledge, we perform the first GWAS of TN. Future studies should be aimed at understanding the extent to which genetic variation stratifies response to neuropathic pain medication and whether genetic information may be used to identify patients who are likely to benefit (or not) from surgical intervention.

## Introduction

Trigeminal neuralgia (TN) is a debilitating neuropathic pain disease characterized by unilateral paroxysmal, lancinating pain along the distribution of 1 or more branches of the trigeminal nerve. Pain can occur stochastically or be triggered by noxious stimuli such as changes in temperature, light touch, or other sensory stimuli.^1^ TN affects approximately 27 out of 100,000 individuals, although the true incidence may be significantly higher depending on the population.^2^ First-line treatment of TN is the sodium channel blocker carbamazepine, based on the hypothesis that neuronal hyperexcitability underlies the pathophysiology of TN.^3, 4^ However, ∼50% of patients do not experience significant symptom reduction with carbamazepine.^5^ Agents that modulate GABA signaling (i.e., gabapentin) have been used, especially in individuals with multiple sclerosis.^6^ When medication fails to result in clinical improvement, surgical treatments such as microvascular decompression (MVD)^7, 8^ or radiosurgery, among others, are performed.^9^ However, population-based anatomical studies have identified many asymptomatic individuals with vascular compression of the trigeminal nerve^10^ and ∼30% of individuals undergoing MVD experience recurrent symptoms,^7^ suggesting that the etiology of at least some portion of individuals with TN is not due to aberrant anatomy. Furthermore, the clinical course of TN patients without radiographic evidence of trigeminal nerve compression may be more severe (i.e., earlier age at clinical presentation).^11^ Overall, these data indicate that additional factors contribute to TN risk.

We hypothesized that, analogous to other neuropathic pain disorders,^12^ there is a substantial genetic component underlying TN risk. Genetic studies of TN have identified 1 single nucleotide polymorphism (SNP) in a serotonin transporter gene in a series of 244 TN patients, loss of function mutations in the GABA_A_ receptor channel gamma-1 subunit (GABRG1), and significant variant burden in the α-1 subunit of Ca_v_3.2 (CACNA1H) in a series of 290 TN patients using whole-exome sequencing.^13, 14^ However, no genome-wide association studies (GWAS) of TN have been performed, significantly impairing our understanding of the genetic basis of the disease. Thus, to expand on these studies, we perform the first GWAS and largest genetic study of TN to date across 1,188 TN cases from three independent biobanks: BioVU,^15^ FinnGen,^16^ and the UK Biobank.^17^ Here we provide evidence for genetic variation conferring significant risk to TN and suggest avenues for incorporating genetic information into clinical care.

## Methods

### Patient cohort description

We used BioVU,^18^ a deidentified health record linked to genetic data, to identify 561 patients with TN and 7,359 control individuals. TN cases in BioVU were first identified using ICD9 (350.1) and ICD10 (G50.0) billing codes before being manually reviewed and selected based on a neurologist or neurosurgeon’s diagnosis in the electronic health record. GWAS summary statistics from FinnGen (G6_TRINEU, 438 cases and 121,360 controls)^16^ and the UK Biobank (189 cases and 361,005 controls)^17^ were also included. Thus, the total cohort consisted of 490,912 individuals (1,188 TN cases and 489,724 controls). Institutional review board approval was provided by Vanderbilt University.

### Genome wide association study analysis

Whole-genome data in BioVU was obtained using the MEGA array and imputed using the 1000 genome reference panel (phase 3). GWAS quality control and imputation were performed as previously described.^18, 19^ Age, sex, and five genotype-based principal components (genomic ancestry) were used as covariates in logistic regression models to identify TN-associated SNPs. We used GWAS summary statistics in the UK Biobank^17^, which had been genotyped using Illumina HumanCoreExome array and imputed using the Haplotype Reference Consortium panel (< 50 cases and 15,582 controls; http://pheweb.sph.umich.edu/pheno/343). SNP association statistics had been calculated using saddlepoint approximation, adjusted for age, sex, and four principal components. Additional information can be found at http://pheweb.sph.umich.edu/pheno/343. We also utilized GWAS summary statistics in FinnGen^16^, which had been genotyped using Illumina and Affymetrix chip arrays and imputed using the SISu v3 imputation reference panel of 3,775 whole genomes. SNP associations had been determined after controlling for age, sex, 10 principal components, and genotype batch as covariates. For additional information on the construction and data elements within the Finngen consortium, see https://finngen.gitbook.io/documentation/.

We performed fixed-effect meta-analysis and appropriate genomic control using METAL.^20^ Genome-wide significance was defined as p < 5.0×10^−8^. Linkage disequilibrium (LD, r^2^) was determined using the LD matrix tool (NCI) leveraging the 1000 genome European ancestry reference dataset.^21^

## Results

We performed a genome-wide association study (GWAS) of TN utilizing whole-genome genetic information from 490,912 individuals (1,188 TN cases and 489,724 controls) across three independent DNA biobanks linked to phenotypic information: BioVU, FinnGen, and the UK Biobank. We identified an intergenic locus on chromosome 1p22.2 flanked by two genes – zinc finger protein 326 (*ZNF326)* and small nucleolar RNA C/D box 36 (*SNORD3G*). This locus contains 5 single nucleotide polymorphisms (SNPs; rs77449572, rs543311093, rs35117749, rs71666259, and rs116010656) reaching genome-wide significance (p < 5.0 x 10^−8^), where rs77449572 is the sentinel variant in the locus (p = 1.72 x 10^−9^) (Figure 1A). As highlighted by the heatmap of r^2^ statistic^22^ values, a number of SNPs in the 1p22.2 locus reaching genome-wide significance with TN are in moderate linkage disequilibrium (LD) (Figure 1B).

**Figure 1.**
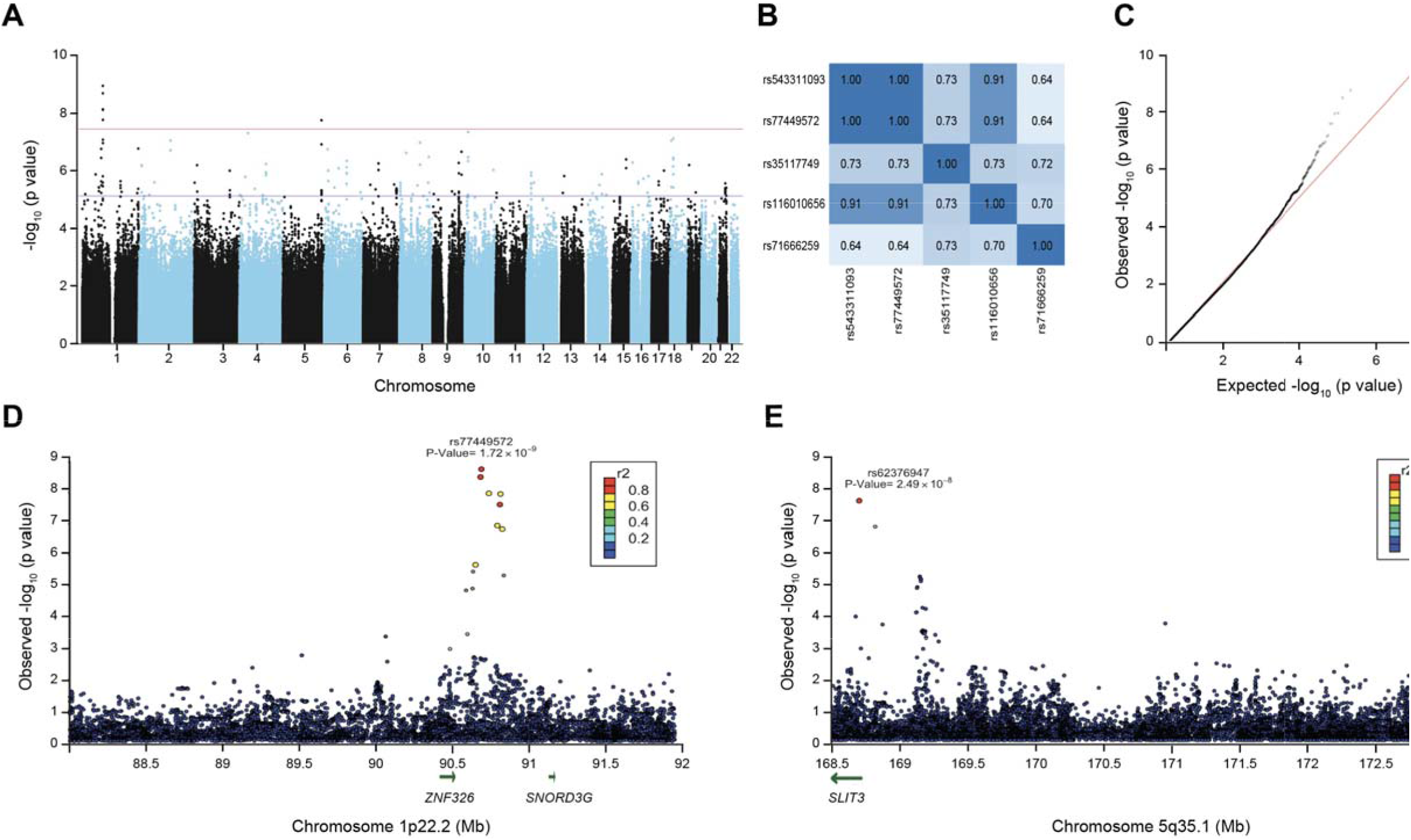
Genome-wide association study of trigeminal neuralgia (TN). (A) Manhattan plot of single nucleotide polymorphisms (SNP) associated with TN (1,188 TG cases and 489,724 controls). (B) Heatmap of r2 values for linkage disequilibrium between SNPs within the locus on chromosome 1 reaching genome-wide significance. (C) Q-Q plot of the distribution of SNP associations with TN. (D) LocusZoom plot of region on chromosome 1p22.2 around the sentinel variant rs77449572 associated with TN. (E) LocusZoom plot of region on chromosome 5q35.1 around the sentinel variant rs62376947 associated with TN.

To infer functional consequences of these variants in biological samples relevant to TN pathophysiology, we utilized Haploreg,^23^ a resource for exploring functional annotations across non-coding regions of the genome in diverse array of cell and tissue types. This approach identified rs77449572 overlapping an enhancer element in cortex derived neurospheres and germinal matrix. In addition, rs71666259 and rs116010656 overlap enhancer elements in embryonic stem cells (HUES48), suggesting that these genetic variants may confer risk to TN by regulating gene expression. We also identified a second locus on chromosome 5q35.1 containing sentinel variant rs62376947in an intronic region of *SLIT3* reaching genome-wide significance (Figure 1A, p = 2.49 x 10^−8^).

Population stratification has been adequately accounted for in our association analysis, as confirmed by a Q-Q plot which demonstrates that there is no global inflation in the test statistic – i.e., no early separation of the expected from the observed (Figure 1C LocusZoom plots show the magnitude of the genome-wide significant associations for the sentinel variants and additional variants in the 1p22.2 (Figure 1D) and the 5q35.1 (Figure 1E) loci. Collectively, these data highlight genetic factors conferring risk to TN.

## Discussion

We perform the first GWAS, to our knowledge, and largest genetic study of TN to date, revealing two independent loci reaching genome-wide significance with TN. We exploit 3 of the largest electronic health records databases linked to genetic information data available and identify two independent genome-wide significant loci. Functional genomics analysis using the Haploreg resource^23^ suggests that variants within the 1p22.2 locus augment gene expression in tissues of neuronal origin. While further molecular studies are needed to determine the precise mechanism by which variation in these loci contribute to TN, this study identifies genetic loci for further experimental testing to understand how genetic variation contributes to the pathophysiology of TN.

*The locus on chromosome* 1p22.2 is located in an intergenic region between *ZNF326* and *SNORD3G. ZNF326* codes for a protein that is a core component of the DBIRD complex that coordinates transcript elongation and alternative splicing at the interface of mRNPs and RNA polymerase II.^24^ In addition, *ZNF326* is highly expressed in the brain and spinal cord (gtexportal.org) and has been shown to play a role in neuronal differentiation, neurodegeneration through regulation of hippocampal volume, and cancer.^25-27^ On the other hand, *SNORD3G* codes for a small nucleolar RNA of largely unknown function. Furthermore, *SLIT3* does not appear to be highly expressed in neuronal tissues (gtexportal.org), but has been shown to be a critical regulator of angiogenesis, monocyte migration, and axon regeneration during peripheral nerve damage.^28-30^ Thus, functional validation of these genes may enable identification of a mechanistic basis for TN pathophysiology and infer development of novel therapeutics.

A number of critical questions remain. First, what is the role of pharmacogenes in regulating response to neuropathic pain medications used to treat TN? Clinical management of TN is challenging, and elucidation of the genetic basis of the disease may inform strategies to understand the role of genetic variation in clinical management.^31^ Given that ∼50% of TN patients do not experience significant symptom reduction with carbamazepine^5^ and ∼30% of individuals with TN undergoing MVD have recurrent symptoms,^7^ it is likely that genetic variation may play a large role, consistent with other neuropathic pain disorders.^32^ Furthermore, it is possible that use of genetic information may assist in identifying appropriate treatment approaches. Specifically, while HLA-B∗1502 is associated with severe adverse effects related to carbamazepine use,^33^ variation in cytochrome p450 genes (specifically, CYP3A4) also regulates metabolism of the drug.^34, 35^ There are likely complex epistatic interactions between these genes, coupled with heretofore unknown rare and common genetic variants influencing carbamazepine (and other neuropathic pain medications) efficacy.

Next, what is the role of genetic variation in defining subtypes of TN? For instance, are patients that present earlier in life, have bilateral symptoms, with no obvious vascular compression of the trigeminal nerve, and/or patients that still experience symptoms post-MVD genetically distinct subtypes? Additional phenotypic refinement and subgroup analysis may reveal a role for genetic risk factors within these populations. For instance, do patients with TN who present earlier harbor large effect-size damaging variants? Further resolution of TN subtypes - stratified by clinical presentation, genetic risk factors, or both – may expand the applicability of genetic information in clinical management. In addition, understanding how genetic variation confers on pharmacologically actionable targets and pathways will improve clinical care. Finally, these data may identify candidates for surgical intervention sooner, therefore limiting the already severe morbidity associated with TN.

## Limitations

While we had access to a deidentified electronic medical record for the subjects in BioVU, we did not have similar phenotyping details for the UK Biobank and Finngen individuals which rely on ICD classifications and summary statistics.^16, 17^ In addition, we were not able to interrogate the contribution of rare (i.e. *de novo* mutations) which have been shown to play a role in TN.^13, 14, 36^ There is likely to be environmental contributions and phenotypic heterogeneity^37^ necessitating evolving approaches to understand the genetic basis of the disease.

## Conclusions

We present results from the first GWAS and largest genetic study of TN to date. These data identify genetic risk factors for TN which will inform our understanding of the genetic basis of the disease and potential avenues for treatment. Future studies should be aimed at understanding the extent to which genetic variation predicts (as part of a polygenic risk score) disease liability across the genome, stratifies response to neuropathic pain medication, and identifies individuals who are likely to benefit from surgical intervention.

## Data Availability

All data will be made available by the corresponding author, Dr. Andrew Hale.

